# Temporal Dynamics of Viral Load and False negative Rate Influence the Levels of Testing Necessary to Combat COVID19 Spread

**DOI:** 10.1101/2020.08.12.20173831

**Authors:** Katherine F. Jarvis, Joshua B. Kelley

## Abstract

Colleges and other organizations are considering testing plans to return to operation as the COVID19 pandemic continues. Pre-symptomatic spread and high false negative rates for testing may make it difficult to stop viral spread. Here, we develop a stochastic agent-based model of COVID19 in a university sized population, considering the dynamics of both viral load and false negative rate of tests on the ability of testing to combat viral spread. Reported dynamics of SARS-CoV-2 can lead to an apparent false negative rate from ~17% to ~48%. Nonuniform distributions of viral load and false negative rate lead to higher requirements for frequency and fraction of population tested in order to bring the apparent Reproduction number (Rt) below 1. Thus, it is important to consider non-uniform dynamics of viral spread and false negative rate in order to model effective testing plans.

## Introduction

As schools consider their return to normal classes, they are relying on the use of tests to combat COVID-19 transmission (Bergstrom 2020). With little information about how COVID-19 will spread through schools, decision-makers are turning to models of viral spread to estimate the amount of testing and the testing frequency required to allow a normal return to schools, as well as other interventions (Bradley, An et al. 2020, Grossman and Peck 2020, Paltiel, Zheng et al. 2020).

Central to the efficacy of mathematical models is the choice of the parameters in those models that describe the spread the disease. In order to model testing, the model must make assumptions about how long after infection a virus is present at a level that can be detected as well as frequency of the false negative rate. Considerations about the rate of transmission of disease are also important because high levels of transmission prior to symptom onset make it harder to control the outbreak (Hellewell, Abbott et al. 2020). Both detection of virus by a PCR based test and transmission of disease to another person are processes that should be proportional to viral load in the patient because the presence of virus in the patient serves as the infectious agent and as the template for the test. Viral load by day after symptom onset has been measured by He et al. and used to estimate the dynamics of viral load prior to symptom onset (He, Lau et al. 2020). They found that virus levels likely start rising just over two days before symptom onset, and that ~44% of transmission may occur prior to symptom onset (He, Lau et al.) (Figure 1).

**Figure 1:**
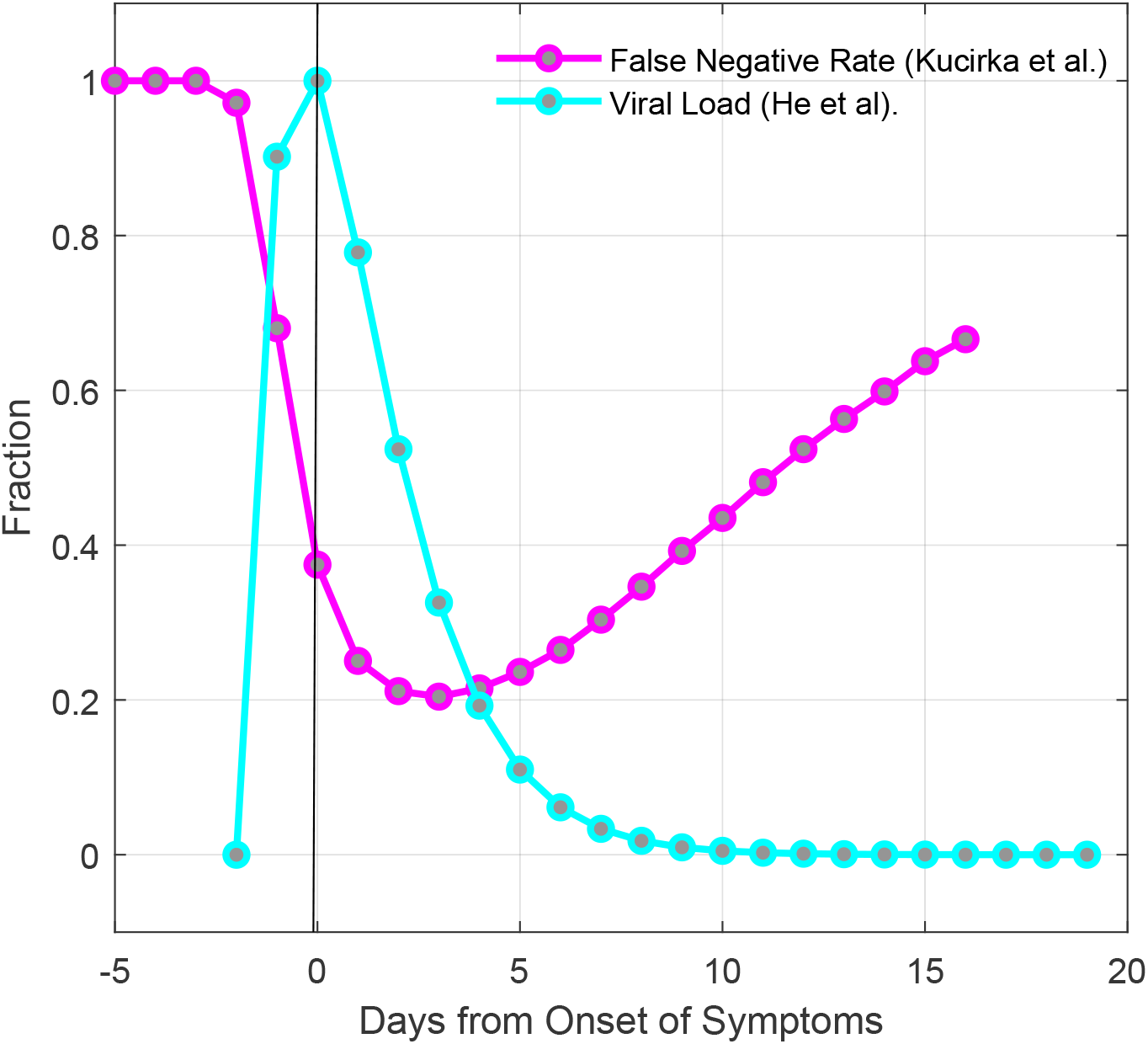
Viral load data and test false negative rate data both suggest that SARS-CoV-2 is undetectable until ~2 days prior to symptom onset. Shown in cyan is the viral load data by day from onset of symptoms from He et al. (He, Lau et al. 2020). Shown in magenta is the false negative rate of tests by day from Kucirka et al. (Kucirka, Lauer et al. 2020). Viral load begins increasing ~2 days before symptom onset, at the same time that the false negative rate of tests begins dropping.

Assessing the efficacy of tests relies upon understanding the false negative rate of testing. False negative rate testing can be broken down into two basic types of false negative, one is a technical failure where the test fails on a sample with detectable levels of virus. Another type is a false negative due to the latent period of the virus, where there is not yet sufficient viral titer in the sample for it to be detected by the test. The viral load data from He et al would suggest that prior to 2.4 days before symptom onset, infected people may not have sufficient virus to be detectable by a test. In a study by Kucirka et al, the dynamics of false negative rate over time was determined by examining data on false negative test in patients who were eventually found to be positive (Kucirka, Lauer et al.). False negative rates were found to be 100% until two days prior to symptom onset and they reached a minimum of approximately 25% two days after symptom onset (Figure 1).

These two studies represent two different data sets that can inform assumptions about viral load, as the ability to transmit disease and detect infection are both likely to be proportional to viral load. While He et al directly measured viral load starting after symptom onset and estimated earlier data points, Kucirka et al. measured the likelihood of a positive test relative to symptom onset and collected data points from presymptomatic patients. The data from both studies predict that detection and viral spread are likely to begin approximately 2 days before symptom onset.

The ability of testing to slow the spread of disease is related to the accuracy and function of the test but also to how fast the disease spreads. In order to stop disease spread, each infected person must, on average, infect less than one other person (an effective Reproduction number (Rt) below 1) (Inglesby 2020). If a large amount of transmissibility occurs in a small window of time, it is more difficult to identify the infected individuals before they transmit to more than one person (Hellewell, Abbott et al. 2020). We hypothesize that the interplay between an undetectable period during incubation and a non-uniform distribution of transmissivity leads to different outcomes for the efficacy of tests in combating disease spread compared to simple estimates of a uniform chance of transmission and a uniform false negative rate. To examine this, we developed a stochastic agent-based Susceptible-Exposed-Infectious-Recovered (SEIR) model of 10,000 students, roughly the size of the University of Maine. We find that the period of undetectable virus leads to a high basal apparent false negative rate, regardless of test sensitivity. When we consider the scenario where only testing is used to combat spread, we find that a simple model that assumes uniform viral spread and perfect tests predicts that testing everyone every 14 days may be sufficient to bring the Rt below 1. However, a model using the combination of disease spread based on the viral load data from He et al. and the dynamic false negative rates for tests from Kucirka et al. predict that as much as 100% of the population may need daily testing to bring the effective R0 below 1 and stop viral spread. While lower levels of testing can be effective in the presence of other interventions such as masking or social distancing, we conclude that the dynamics of an undetectable period, viral transmission that is biased early in the disease, and dynamic false negative rates significantly change the predictions of an SEIR model, and these factors should be considered when developing models to plan for public health interventions to combat COVID19.

## Methods

### Model

We chose to build a stochastic agent-based model for two reasons: 1) it would allow us to easily implement nonuniform probabilities over the course of infection and 2) a stochastic model would capture the inherent noise in a system that is presumed to start with a small number of infected cases. We implemented the model in MATLAB using the indicated probabilities and if-then statements. The test was performed with 10,000 individuals to represent the college student body. The model runs daily for 120 days, approximating a semester. The basic structure of the model is outlined in figure 2. Because it is a stochastic model, we perform 100 independent runs (Figure 3), and report the median and 95^th^ percentile results. The model can be found on GitHub at https://github.com/Kelley-Lab-Computational-Biology/coronamodel.

**Figure 2:**
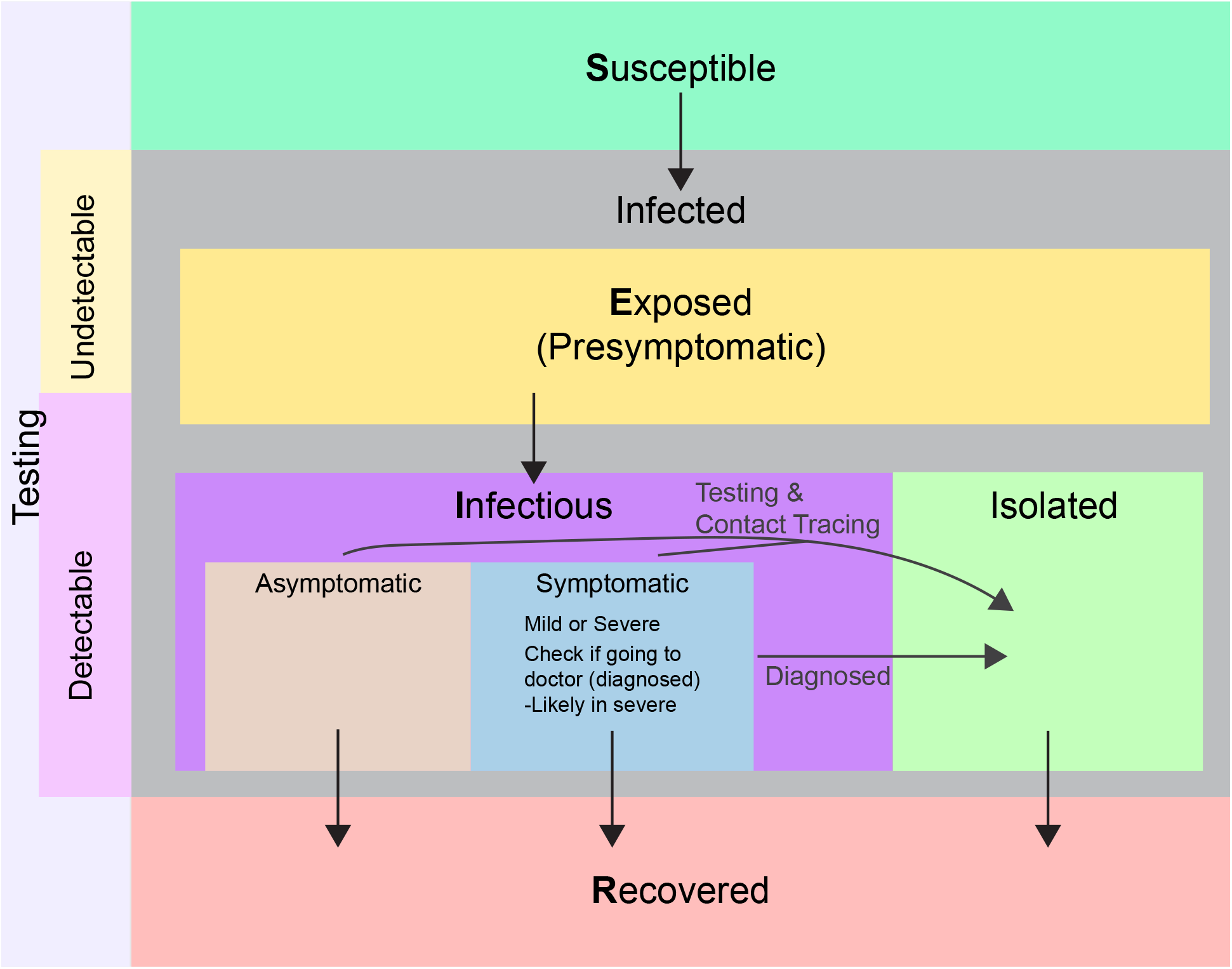
Diagram of the stochastic agent-based model. This is a stochastic SEIR model implemented in MATLAB. Each transition in state is based on if-then statements with specific probabilities described in figure 4. Individuals start as susceptible, and the initial population is seeded with 10 random infected individuals, each starting at a random point of progression through the disease, and with random symptoms. Upon being infected, an individual become exposed (presymptomatic), and is assigned a day for symptom onset. Detectability for testing and infectiousness both begin at 2 days prior to onset of symptoms. Infectious individuals can be either asymptomatic, or symptomatic with mild or severe symptoms. Those with severe symptoms will self-isolate and initiate contact tracing through seeking medical attention. Asymptomatic individuals and those with mild symptoms can be isolated through contact tracing or through detection by a test. Infectious individuals will recover randomly with a median time of 14 days.

**Figure 3:**
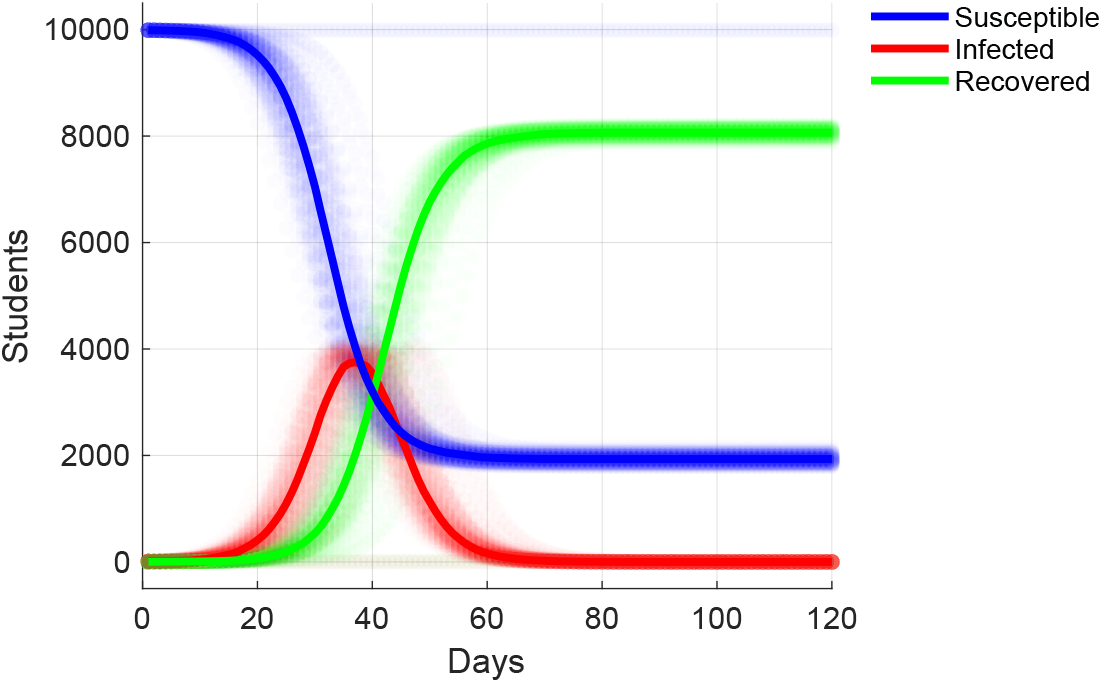
Example of 100 independent simulations with the model. Shown are susceptible, infected (encompassing exposed, infectious, and isolated individuals), and recovered individuals in simulations where no interventions were implemented. Each individual simulation is represented as semi-transparent points, while the median value of all simulations is plotted as a line.

### Symptoms

For the timing of symptom onset, we used the symptom onset distribution calculated by He et al. This distribution has a median onset time of 4.2 days, and 99% of cases experience symptom onset by 14 days (Figure 4A) (He, Lau et al. 2020).

**Figure 4:**
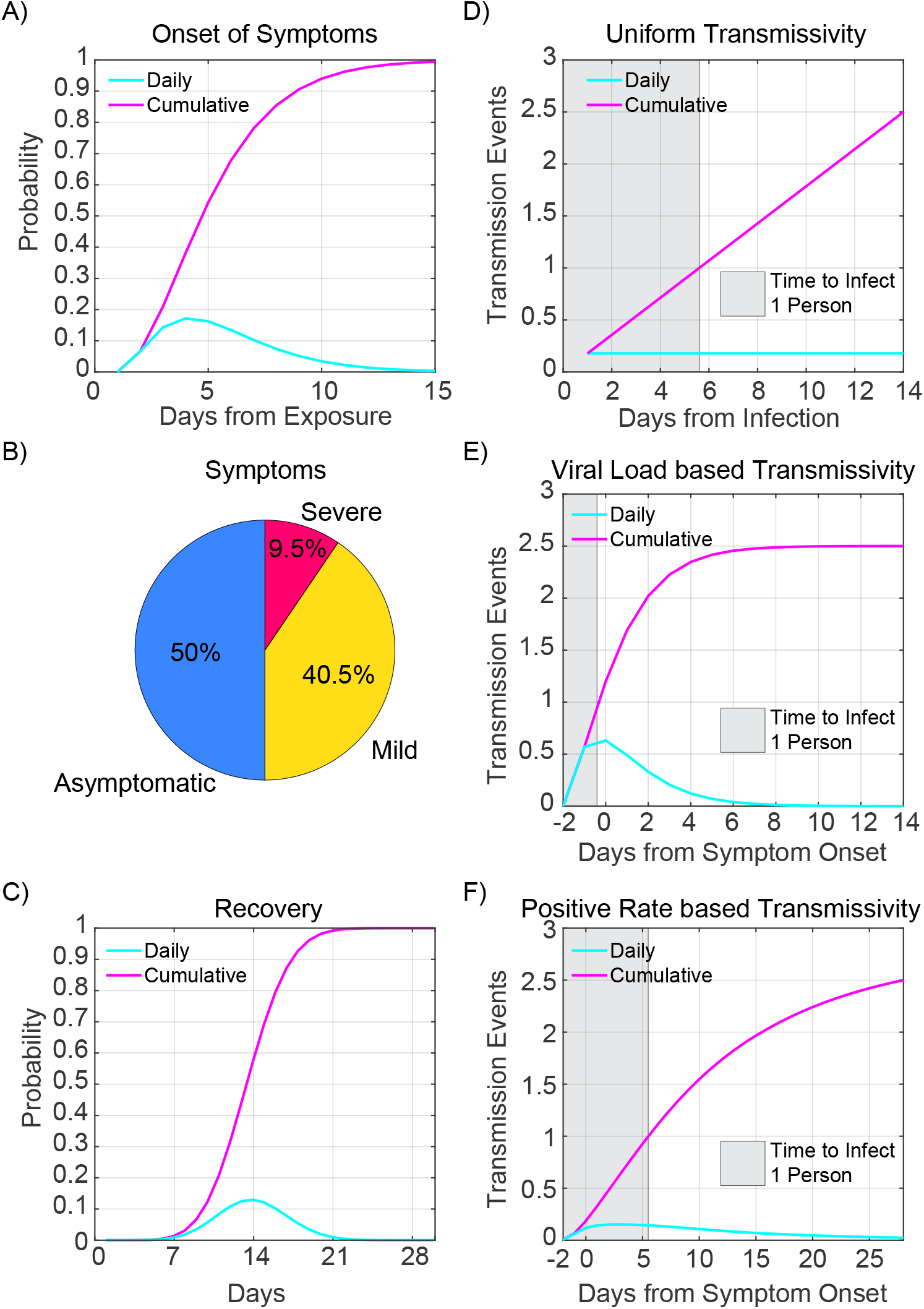
Model Parameters. A) Probability distribution of onset of symptoms from He et al. B) Breakdown of symptom groups in the model. C) Probability distribution of recovery based on a median time to recovery of 14 days. D) R0 of 2.5 scaled to a uniform transmission probability distribution. The gray box indicates where the cumulative probability reaches 1. Individuals must be detected prior to this, on average, in order to reduce the apparent R0 below 1. E) The R0 of 2.5 scaled to the viral load based on He et al. The gray box is the same as above. D) The R0 of 2.5 scaled to the positive test rate from Kucirka et al. This was done because the changes in positive test rate are likely related to viral load, and so may be an alternative representation of transmission likelihood. The gray box is the same as above.

The CDC reports an overall asymptomatic rate of 40% (CDC 2020), but we are concerned about the likely asymptomatic rate among a young population. When the aircraft carrier Theodore Roosevelt had an outbreak of COVID-19, they reported that as many as 350 out of 600 sailors were asymptomatic for an asymptomatic rate of 58% (Correll 2020). As the population aboard a navy ship are likely to skew younger and healthier than the population as a whole, we felt they may be more representative of college age students. Thus, we assumed an asymptomatic rate of 50%. People who are symptomatic are then assigned either mild or severe symptoms, based on CDC data that 81% of people experience mild symptoms, 14% severe, and 5% critical (CDC 2020). We consider severe and critical together, as we expect both to seek medical assistance, and then be isolated from the general population. We also assumed that these number represent the percentages of symptomatic people, so ultimately the model assigns 50% asymptomatic, 40.5% mild, and 9.5% severe (Figure 4B). We assume that those experiencing severe symptoms seek medical attention at the beginning of symptom onset and are isolated, and initiate contact tracing. For this paper, we assume that mild cases do not self-isolate, as they may not realize that their symptoms are COVID-19 related, or they may be reluctant to identify themselves as ill for fear of isolation and removal from their normal college activities (Pagoto 2020). While this assumption will make the spread of the disease harder to contain, we did not want to make the model unduly optimistic about behavior.

### Recovery

The CDC reports median recovery time as 14 days for mild illness (CDC 2020). We assume the same recovery period for asymptomatic people. Because severe illness results in medical attention and isolation, we did not consider the extended recovery period for severe illness as it would not change transmission in our model. The recovery probability distribution is modeled as a binomial distribution with a mean of 14 days (Figure 4C).

### Probability of viral spread

The model assumes an R0 of 2.5 (CDC 2020). Each individual in the model receives an R0 normally distributed around 2.5 to allow for variability in transmissibility between people. We took three different approaches to viral transmission probability. 1) We assume a uniform daily transmission probability equal to 2.5/14 (R0 / median time of illness) (Figure 4D). 2) We assume that daily transmission rate is proportional to viral load, and so we scale the R0 to the viral load data from He et al. (He, Lau et al.), where transmission starts 2 days prior to symptom onset (Figure 4E). 3) A daily transmission probability scaled to the false negative test rate reported by Kucirka et al. (Kucirka, Lauer et al. 2020), under the assumption that the dynamics of the false negative rate are related to the viral load (Figure 4F). For each of these assumptions about viral spread, people must be detected on average before they spread virus to one other person on average (R0 below 1). We have indicated in Figure 4 D, E, and F with a shaded rectangle the time in which sick individual must be detected to keep the average number of new infections below 1.

### Testing

Tests can be administered to the entire population, or to randomly selected subsets of the population either daily or at varied frequencies. For the purposes of this study, we assumed tests are resolved on the day they are administered. We consider a few scenarios for false negative rates: 1) Perfect tests, where there is no chance of a false negative rate, and there is no period of undetectable infections. 2) Our “simple” scenario where the virus is undetectable until 2 days prior to symptom onset, after which tests have a uniform 5% false negative rate. 3) Dynamic false negative rates based on those measured by Kucirka et al (Kucirka, Lauer et al.). Like the simple scenario, there is no chance of detecting an infected individual prior to 2 days before symptom onset. We do not consider the ramifications of false positive rate. While the false positive rate is important due to the burden that incorrectly identified cases place on resources (Paltiel, Zheng et al. 2020), that consideration does not affect the Rt of the system.

### Contact tracing

For each individual in the model, we store the identity of the source of their infection, and the identities of people they transmit to. If someone is identified as sick by self-isolating and seeking medical attention, or if they are identified by a randomly administered test, contact tracing is initiated. We assume a 75% chance to identify each contact of the individual.

## Results

### Nonuniform false negative rates can delay detection of infected individuals

Why are we concerned about uniform versus nonuniform false negative rate? To illustrate the issue, we can examine the first day of disease progression at which an infected individual is likely to be detected when tested daily (Figure 5). We compare three different false negative rate dynamics over 14 days of disease progression assuming testing every day, and we assume symptom onset at day 5. The average false negative rate of each is the same (50.42%), but the way the rates change over time differs. We have indicated with a gray rectangle the two days prior to symptom onset that may represent as much as 44% of viral transmission capability (He, Lau et al. 2020). 1) A completely uniform false negative rate leads to most infected people being detected by day 3, prior to becoming infectious. 2) An undetectable period followed by a uniform rate of detection catches most individuals by day 5 (it is, after all, just a two day offset of (1), with the uniform rate rescaled to still average to the same overall false negative rate). These assumptions about the dynamics of viral spread allow more people to spend time in the infectious period prior to being detected than the completely uniform assumption. 3) The dynamic false negative rates of Kucirka et al. means that few individuals are likely to be caught prior to the potential for significant viral spread (Kucirka, Lauer et al. 2020).

**Figure 5:**
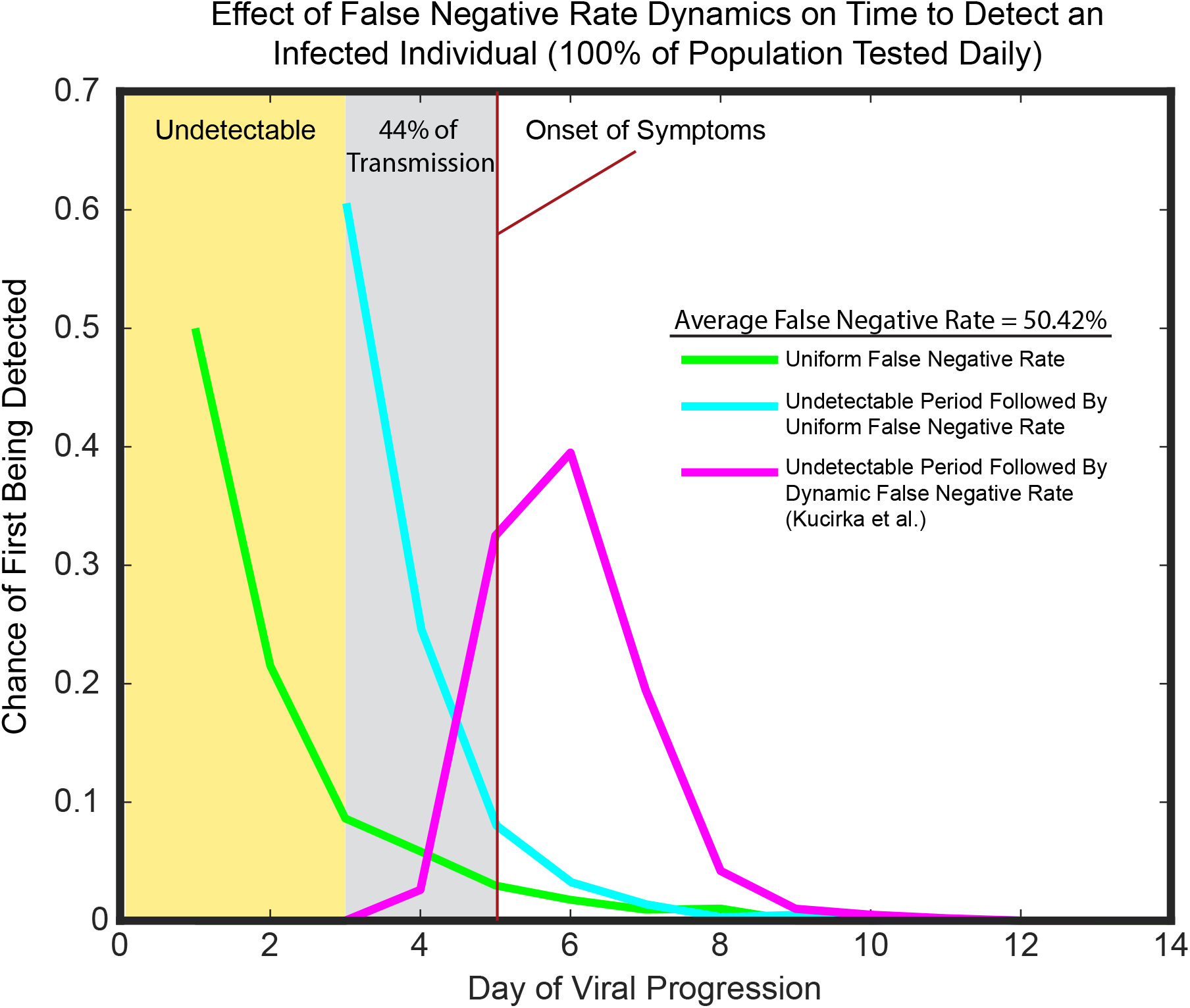
Non-uniform false negative dynamics can delay detection of infected individuals. Shown is the chance of first being detected at each day of disease progression based on three scenarios with the same average false negative rate across the 14 days shown, but different temporal dynamics. For this graph, we assume that symptom onset begins at day 5. In yellow is the undetectable period prior to 2 days before symptom onset. The two days before symptom onset is shown in gray. Viral load data suggests that as much as 44% of transmissibility may occur in these two days. The data is histograms of the first day that an individual would be detected by a daily test with the given false negative rate dynamics.

### An undetectable period leads to high apparent false negative rates

The viral load data from He et al. suggests that virus first rises to a detectable level two days prior to symptom onset. Since viral RNA is the template for PCR based tests, the ability of the tests to detect the virus will be dependent upon the viral load, so we made the simple assumption that virus was undetectable prior to 2 days before symptom onset, and that it was uniformly detectable after this point. The measured false negative rates reported by Kucirka et al. validate this assumption, and provide daily false negative rates after viral load begins increasing. We made a separate model using the Kucirka et al. measured false negative rates.

We used the model to test the effect of these different assumptions on the overall false negative rate that would be encountered during random testing for the virus, where the people who are positive are randomly distributed through the progression of the disease. For example, while the median of symptom onset is between 4 and 5 days, 12% of cases would have a start of symptoms at 9 days or later. In this case, there would be at least 7 days during which there is insufficient virus present to detect an infection, regardless of the efficacy of the test. Simulations were run 100 times, and the median value of the false negative rate is reported (Figure 6). We found that the simple model, which assumes uniformly perfect tests after 2 days prior to symptom onset displays an apparent false negative rate of 17%. In the case of the Kucirka data, which has both the undetectable time period before virus replication begins and the measured daily false negative rates afterward, which reach a minimum of ~25% two days after symptom onset, the overall false negative rate is of the simulation was 48%. It is worth reiterating that this is the false negative rate one would experience testing a random group of people, not the false negative rate expected for directed testing, such as testing someone who is symptomatic. The Kucirka et al. false negative data is a compilation of both the false negative rate of the test, and the false negative rate due to the viral infection dynamics. The simple model considers only the false negative rate from the viral dynamics and places the lower bound at 17% false negative, which is large but within the realm of consideration (Paltiel, Zheng et al. 2020).

**Figure 6:**
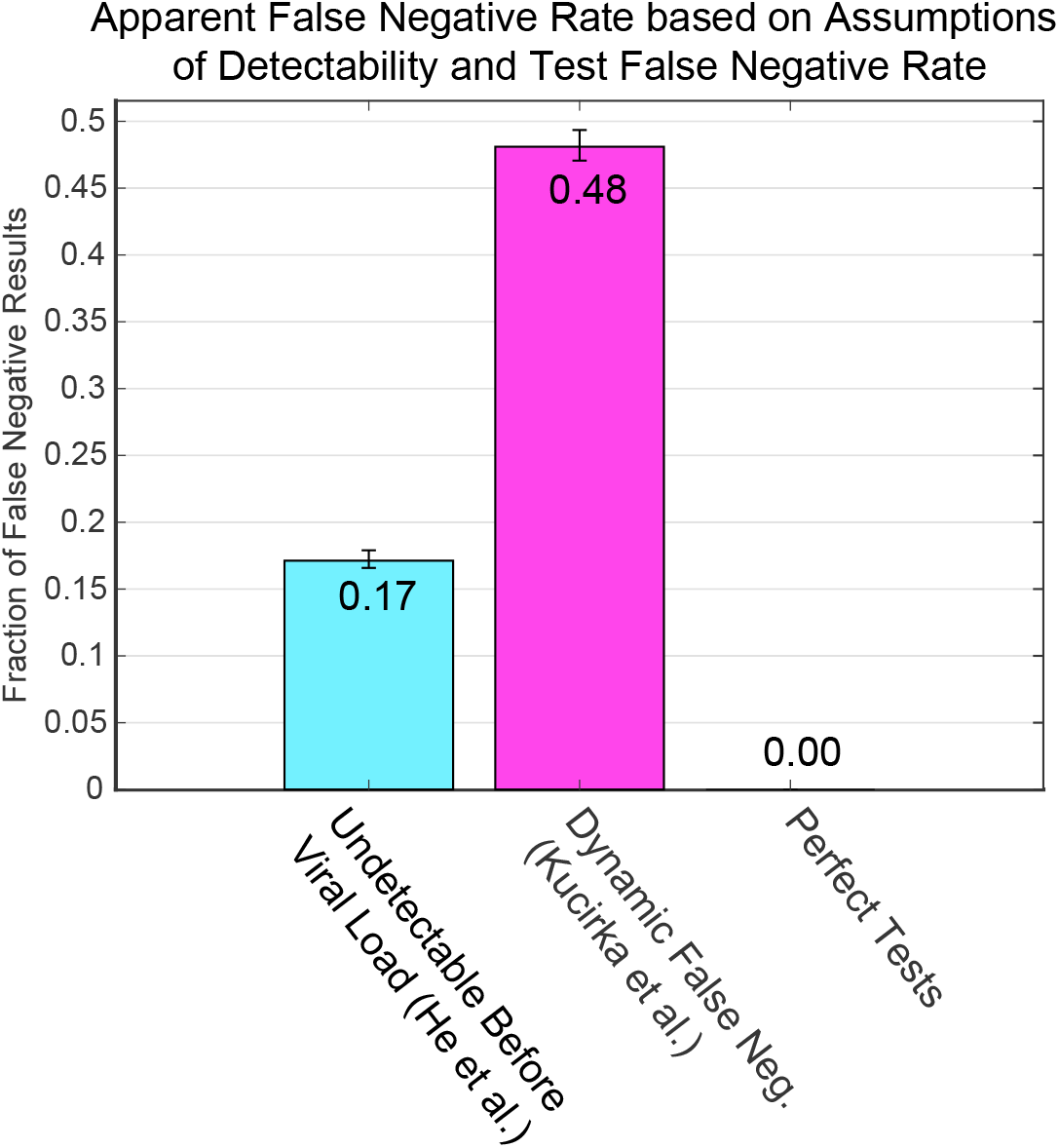
The undetectable period and temporal dynamics of the false negative rate lead to high apparent false negative rates. In cyan is shown the model run with the simple assumption that infected individuals were undetectable before viral load begins, based on the He et al. data., and that after that point the tests will always detect infected individuals. In magenta, the model uses the dynamic false negative rates from Kucirka et al., in which both test error and inability to detect due to low viral load are mixed together. Also included is the effect of perfect tests.

### An undetectable period and high early transmission levels lead to a need for higher levels of testing

If the effective false negative rate ranges from 17% to as high as 48%, it is likely to affect the level of testing required to combat the spread of COVID-19. We set out to examine the effect of testing on the spread of disease by calculating the effective R0 of the virus when different testing regimens are used, while varying the dynamics of detectability and test false negative rate. We varied the fraction of the population being tested and the frequency of the test for four scenarios. Those scenarios are:

**Scenario 1: “Perfect tests, Uniform Spread”** where we assume no period of undetectability, no false negative rate, and a uniform chance of transmission equal to 2.5 / 14. **Scenario 2: “Simple Undetectability, Fast Spread”** where we assume that the virus is not detectable until 2 days prior to symptom onset, and then has a 5% false negative rate after that point (this 5% false negative rate is a change from the simple assumption above (Figure 6), which assumed perfect tests). This condition uses the He et al. viral load data to scale the R0 (Figures 1 and 4E), which results in ~45% of transmissivity prior to symptom onset. **Scenario 3: “Dynamic False negative, Slow Spread.”** This uses the day-by-day false negative rates reported by Kucirka et al for testing (Figure 1). For transmissivity, we use the day-by-day positive rates from the Kucirka et al data as a stand-in for viral load (Figure 4F). The shape of this profile still biases spread early in the disease, but not as early as the He et al. viral load data. **Scenario 4: “Dynamic False negative, Fast Spread.”** This scenario uses the day-by-day false negative rates from Kucirka et al. for testing, and the He et al. viral load data to scale transmissivity.

These simulations are run with testing being the only intervention being used to combat viral spread. We report the median effective Rt as well as the 95^th^ percentile Rt for each condition because testing regimens that work only half the time may not be useful when considering public health. We see that perfect tests can be effective while testing as little as 25% of the populace every other day (Figure 7). All simulations that do not assume perfect tests require a larger proportion of the population to be sampled under these conditions. Scenario 2 and Scenario 3 result in remarkably similar results for which testing regimens are required for suppression of viral spread. The fast viral-spread and sensitive tests of Scenario 2 are therefore compensated for by the slower viral spread and insensitive testing of Scenario 3. With scenario 4, where the transmission occurs early in the disease and false negative rates are high, only testing of every individual every day was able to bring the Rt below 1. Thus, viral transmission that is biased early in the progression of the disease and higher false negative rates require a more aggressive testing regimen than would be suggested by uniform assumptions.

**Figure 7:**
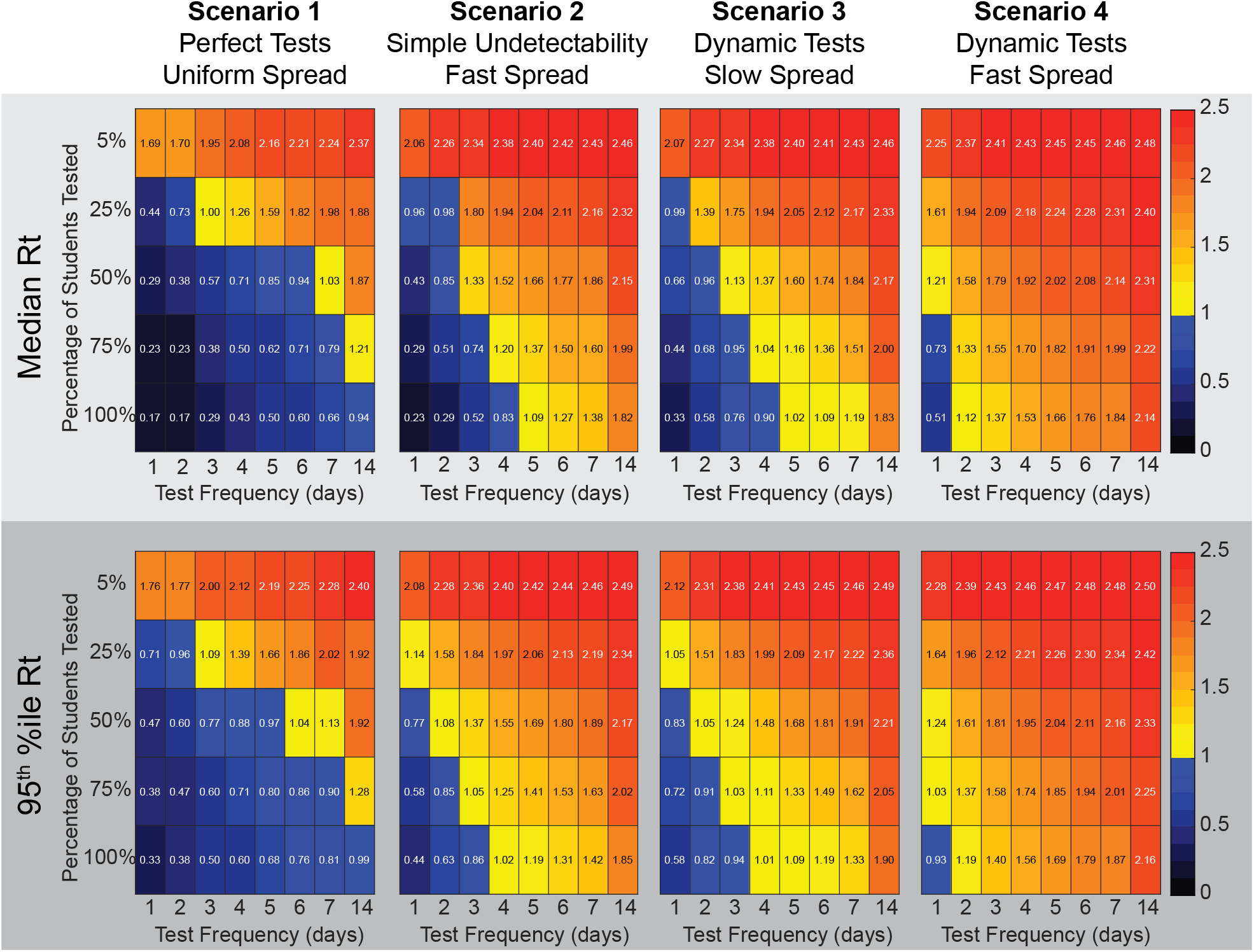
High asymptomatic transmission and dynamic false discovery rate lead to a requirement for more testing to bring the viral spread under control. Heatmaps show the effective Reproduction number (Rt) from 100 simulations run with the given proportion of the population tested at the indicated frequency. The top row of matrices shows the median Rt, while the bottom row of matrices shows the value of the upper 95^th^ percentile. While the scenario 1 perfect tests suggest testing the entire population every two weeks may work to stop spread of the virus, using scenario 4 parameters predicts that testing the entire population daily was necessary.

While these simulations suggest that testing would have be very aggressive to bring viral spread under control, they are not assuming any other interventions. In reality, testing is likely to be component of a multi-pronged approach to combating viral spread. We decided to examine the efficacy of testing under a situation where other interventions had brought the viral spread down, but not below an Rt of 1. A recent study of mask efficacy suggests that surgical or cloth mask wearing can reduce the risk of contracting COVID19 to 33% the risk of those not wearing masks (Chu, Akl et al.). Interestingly, this is similar to the percent decrease in particulates that has been described for a cloth mask (van der Sande, Teunis et al. 2008)(average reduction in particulates to ~31% of control over a 3 hour experiment). We implemented a model where 70% of the population uses masks that reduce transmission rate by 67%. This results in a median apparent Rt of 1.3, and a 95^th^ percentile value of 1.44. We then performed the simulations using the array of testing regimens as above. For this analysis, we used the Scenario 4 conditions of dynamic false negative rate (Kucirka, Lauer et al.) and high early viral transmission dynamics (He, Lau et al.), as these conditions are the hardest to reduce and will give the most conservative results for frequency and amount of testing. We find that under these conditions it would now be possible to bring the Rt below 1 in 95% of cases by testing 25% of the population every day (Figure 8). Testing every person would now be effective when done once a week.

**Figure 8:**
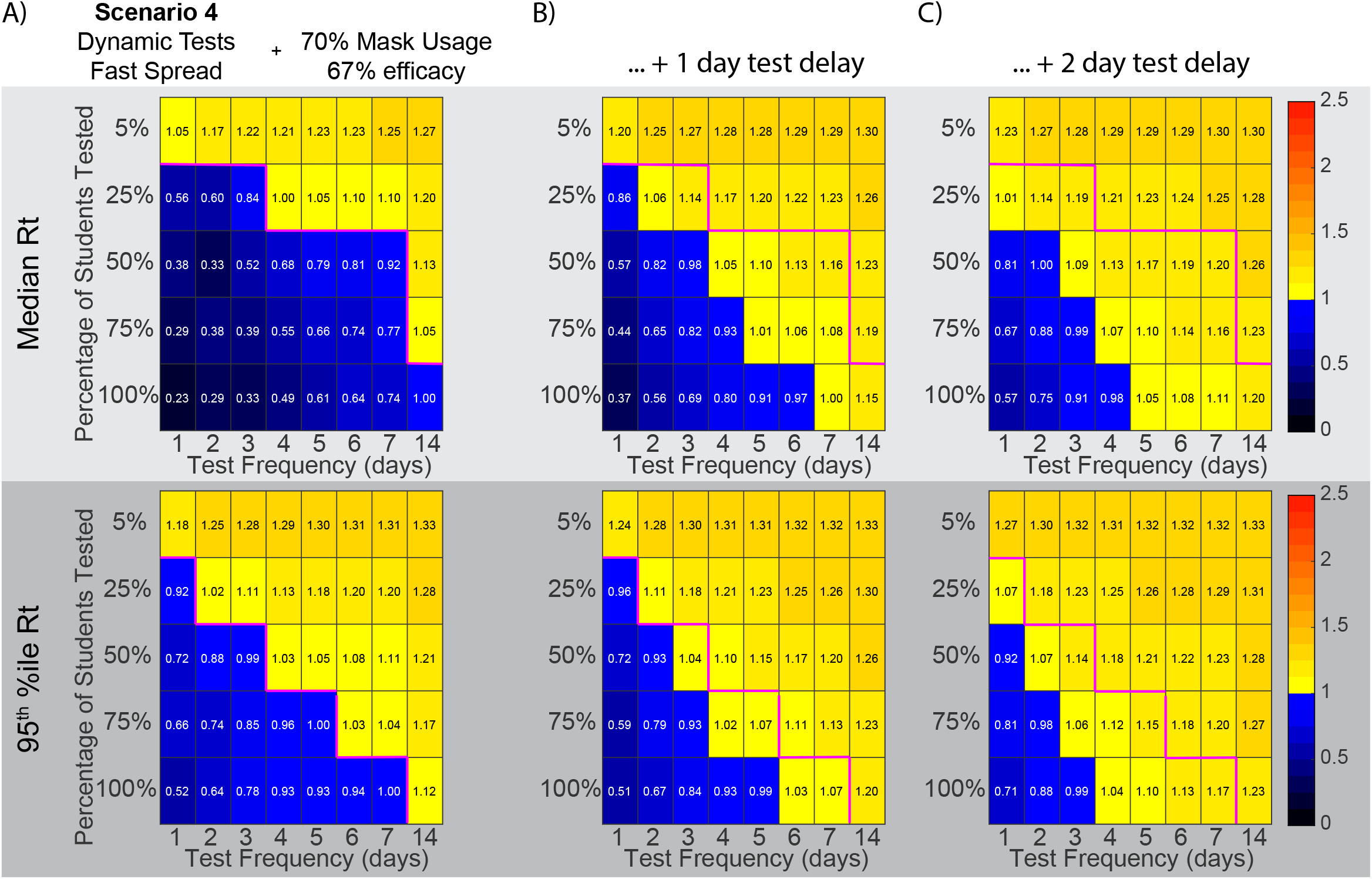
In the presence of masking, fewer tests and lower frequencies of testing can be successful in driving R0 below 1. A) Here we implemented 70% of the population using a mask that is 67% effective with the parameters of Scenario 4, early transmission of virus based on the He et al. viral load data, and dynamic false negative rates for tests based on Kucirka et al. The top row of matrices shows the median effective Reproduction number (Rt), while the bottom row of matrices shows the value of the upper 95^th^ percentile. Masking drove the median Rt from 2.5 to ~1.3. Tests were then able to drive the 95^th^ percentile Rt below 1 with less aggressive testing schemes than in Figure 7. B) The same conditions as (A), with an included 1 day turn around delay in testing results. The magenta line shows the border between an Rt above 1 and an Rt below 1 without a delay. C) As in (B) with a 2 day turn around delay in testing results.

These previous simulations assume instantaneous turnaround time for the test results. Unfortunately, test results may take a day to several days for results to be available. The delay is typically due to the backlog of samples needed to be tested, lack of testing equipment, and the relatively small number of labs and technicians with proper certification (Barone 2020). To analyze the effect on delay in receiving the results, we used Scenario 4 conditions with 70% of the population using masks then performed the simulations using the array of testing regimens as above. In the model, students who are tested and found positive start their isolation after they find out their results along with people isolated due to contact tracing. Figure 8B shows how implementing a one-day delay has a detrimental effect on the testing requirement in order to prevent an outbreak. A one-day delay in receiving test results leads to a requirement for a two-day increase in frequency, as testing the whole population every 5 days would prevent an outbreak compared to testing every 7 days with no delay. Similarly, a two-day delay in receiving test results leads to a four-day increase in testing frequency necessary to prevent an outbreak (Figure 8C). The delay in test results significantly changes the testing frequency requirements in order to prevent an outbreak.

## Discussion

Available data on SARS-COV2 viral load over time and on false negative rate of tests over time both suggest that virus may not be detectable prior to ~2 days before symptom onset, and transmissibility of the virus is biased towards the beginning of disease progression. Here we have examined the effect of nonuniform viral transmission and nonuniform detectability of disease on the efficacy of testing as a means to stop viral spread. We find that the combination of the non-uniform transmission dynamics and false negative rate predict that tests must cover more of the population and be given more frequently than predicted by a model that assumes uniform distributions. Thus, models that make simple assumptions about viral spread, and false negative rate or underestimate the effect of the undetectable period on the apparent false negative rate may recommend less testing than is necessary to stop viral spread.

The parameters used for these simulations (viral load dynamics, false negative rate, efficacy of masks and level of compliance with masking) are not concrete, and are likely to vary between institutions, populations, or areas. As the model parameters approach containment of viral spread, the prevalence of virus in the surrounding community, or other sources of introduction into the system will be more important to the considerations for testing amount and frequency as well as quick turnaround time of results. As such, these results should not be seen as recommendations on specific testing strategies, although the results for Scenario 4 are clearly conservative. Similarly, these studies should not be construed as saying that tests do not work or that tests should not be a part of the public health strategy for combating viral spread. Instead, the takeaway message is that modeling of tests should be done with consideration of the potential for an undetectable period, nonuniform transmission dynamics, and the potential for viral load to influence false negative rate. Each of these considerations alters the conclusions that a model will come to about the number and frequency of tests required to combat viral spread.

There are many reports in the news media of organizations using a negative test result as a prerequisite for engagement in some activity, such as returning to college or attending a summer camp. The Kucirka et al. data on dynamic false negative rate should already give pause to these types of plans, but we show here that testing a population of people who may have a random distribution of progression through disease may have a false negative rate as high as 48%. The possibility of missing ~1/2 of positive individuals by performing a complete testing of the population of interest should be considered when making these plans. This high false negative rate is specific to tests which are performed on a population likely to have a random distribution of viral progression. In situations where the tests are being given because of symptoms, or because of contact tracing, the population being tested would be biased towards later days in the progression of the disease, and the overall false positive rate would be lower than the 48% value. However, even if one were using a test that was 100% sensitive and specific given a sample that contains template, it is likely that they would still experience the ~17% false negative rate due to the latent period of the virus before it begins replicating. Thus, plans to allow people to participate in activities dependent upon a negative test should be aware of the greater than 1 in 6 likelihood of missing an infected person in their testing.

In conclusion, many people are resorting to modeling of disease transmission to assist in the formulation of public health plans for the return to schools and economic activities. When designing these models, simple assumptions of uniformity of transmission and uniformity of false negative rate can give overly optimistic views of the efficacy of testing. These nonuniform dynamics are complicated to implement in a deterministic ODE model, but easier to implement in a stochastic agent-based model. The stochastic model, however, is slow compared to an ODE model. Answering questions about tests does not, however, require a population to be so large as to be unmanageable with a stochastic model, as the trends in testing efficacy should remain the same. Thus, we recommend that stochastic models be used to model efficacy of tests so that complex dynamics can be readily accounted for. The results of stochastic models could then be used to parameterize deterministic models for other uses.

## Data Availability

data and matlab scripts necessary to reproduce this work are available at GitHub

https://github.com/Kelley-Lab-Computational-Biology/coronamodel

## Acknowledgements

We would like to thank Melissa Maginnis, Rob Wheeler, Sara Huston, Joshy George, Yi Li, and Krishna Karuturi for helpful discussions.Contributions: KFJ developed the model, performed the simulations, performed data analysis and assisted with writing the manuscript. JBK conceived of the study, developed the model and wrote the manuscript.

Funding: Work supported by NIH R15GM128026 to JBK

## Notes

### Competing Interest Statement

The authors have declared no competing interest.

### Funding Statement

Work supported by NIH R15 GM128026 to JBK

### Author Declarations

IRB approval not needed.

### Summary of Updates

Added assessment of the effect of test result delay on the efficacy of testing.

